# The Peripheral Use of Low-dose Vasopressors for Safety and Efficacy (PULSE) in the intensive care unit: a prospective, unblinded feasibility study protocol

**DOI:** 10.64898/2026.04.13.26349750

**Authors:** Jessica Flora Wiseman, Stephanie Sibley, Santiago Perez Patrigeon, Maikel Mekhaeil, Michaela Hanley, Miranda Hunt, Tracy Boyd, Brianna Grant, J. Gordon Boyd

**Author notes:** **Contact Information:** Jessica Wiseman, Department of Critical Care Medicine, Queen’s University/Kingston General Hospital, 76 Stuart Street, Kingston, Ontario, K7L 2V7. **Contributors:** JGB and JFW designed the study and wrote the study protocol. JFW drafted and revised the manuscript. SS, SPP, MM, MH, MH, TB, BG, and JGB reviewed and revised the manuscript. JGB is responsible for the overall content as guarantor. **Funding:** This research received no specific grant from any funding agency in the public, commercial or not-for-profit sectors. **Competing interests:** none declared.

## Abstract

**Introduction:** There is increasing interest in the peripheral administration of vasopressors for two main reasons: (1) to expedite vasopressor initiation in patients with refractory shock and (2) to avoid the potential complications associated with central venous catheter placement. The current evidence on the use of peripheral vasopressor administration is primarily based on single-center observational studies. There are inconsistencies in the administration of peripheral vasopressors, including catheter gauge and location, monitoring practices, vasopressor concentrations, and duration of use. This has made it difficult for institutions to develop best practice guidelines. A randomized controlled trial is needed to address this knowledge gap.

**Methods and analysis:** The **P**eripheral **U**se of **L**ow-dose Vasopressors for **S**afety and **E**fficacy (PULSE) in the intensive care unit is a prospective, unblinded feasibility study. Eligible patients will be 18 years or older, have no existing central venous catheter or peripherally inserted central catheter and have the presence of shock requiring a minimum vasopressor dose of any of the following: norepinephrine 0.0625 mcg/kg/min, phenylephrine 0.625 mcg/kg/min, and epinephrine 0.0625 mcg/kg/min. Fifty patients will be randomized 1:1 into either the peripheral venous catheter or central venous catheter group. The primary outcome is feasibility, defined as (1) a recruitment rate of 4 participants per month, (2) a data capture rate of ≥90%, and (3) a <50% conversion rate from peripheral to central access. The secondary outcomes include the safety of peripheral vasopressor use, alive and central-line-free days, the number of attempts needed to place a catheter, volume status, in-hospital mortality rate, ICU and hospital length of stay, and patient-centred important outcomes.

**Implications:** The data collected from this study will inform the design of a definitive randomized controlled trial to assess the safety and efficacy of protocol-driven peripheral vasopressor administration.

**Ethics and dissemination:** This study received approval (6042888) from the Queen’s University Health Sciences/Affiliated Teaching Hospitals Research Ethics Boards. Results of this study will be presented at critical care conferences and submitted for publication.

**Trial registration number:** NCT06920173 (https://clinicaltrials.gov/study/NCT06920173).

## INTRODUCTION

In the intensive care unit (ICU), vasopressors are administered when patients cannot maintain an adequate blood pressure in the presence of shock, which is a life-threatening condition characterized by inadequate tissue perfusion. Historically, the central venous catheter (CVC) or central line has been the preferred route of administration for vasopressors, as prior data have suggested an increased risk of vasopressor extravasation through peripheral venous catheters (PVCs). ^1^When norepinephrine was first used clinically in the 1950s, extravasation rates following peripheral administration ranged from 46% to 60%. ^2^ However, advances made in venous catheter access have significantly reduced this risk, with more recent extravasation rates ranging from 1.8% to 3.4%. ^1,3^

The placement of a CVC requires a skilled clinician and, oftentimes, ultrasound guidance to prevent complications such as pneumothorax, bleeding, or inadvertent arterial cannulation. Complications that may arise during CVC maintenance include deep vein thrombosis (DVT) and central line-associated bloodstream infections (CLABSIs). These recognized complications may occur in more than 15% of patients^4,5^, which is one reason for the growing interest in peripheral vasopressor administration. ^6^Research shows that many clinicians initiate peripheral vasopressors in patients presenting with refractory shock^7-9^; however, most patients will eventually have a central line placed.^7^

## BACKGROUND AND RATIONALE

The current data supporting the use of peripheral venous catheters for administering vasopressors are primarily based on single-center observational studies. Inconsistent monitoring of peripheral lines and variations in vasopressor concentrations, doses, and duration of use may hinder the widespread practice of peripheral administration of vasopressors. ^10-14^Additionally, there are no universal guidelines for the transition threshold for central-line placement, with only weak recommendations from the Surviving Sepsis Guidelines suggesting that vasopressors should be infused through a central line as soon as possible, if resources are available.^15^ Therefore, some clinicians recommend viewing PVCs as a bridge to central access until more reliable evidence becomes available. ^16^A randomized controlled trial is needed to provide high-quality evidence to assess the safety of peripheral administration of vasopressors. Prior to embarking on a definitive trial, a feasibility study is required to inform its design.

### STUDY OBJECTIVES

#### Primary Objective

To determine the feasibility of a randomized controlled trial for the implementation of a peripheral vasopressor protocol. The following factors will be assessed:

1. Recruitment rate (measured as the proportion of eligible patients randomized into the study).
2. Data capture rate (measured as the percentage of data elements obtained for each participant).
3. Conversion rate to central access (measured as the number of participants recruited into the PVC arm who require CVC placement).

#### Secondary Objectives

1. Assess the safety of peripheral vasopressors.
2. Assess alive and central-line-free days.
3. Assess the number of attempts in the placement of the peripheral venous catheter and the central venous catheter attempts.
4. Assess the rate of complications.
5. Determine the amount of vasopressor volume.
6. Assess the 30-day mortality rate.
7. Compare ICU and hospital length of stay.
8. Assess patient-centred important outcomes.

## METHODS AND ANALYSIS

### Design

PULSE is a prospective, unblinded feasibility study to be conducted at a tertiary academic center. It will be unblinded because a central line will be easily noticeable. The study is registered with clinicaltrials.gov (trial registration number: NCT06920173). The trial schema is illustrated in Figure 1.

**Figure 1.**
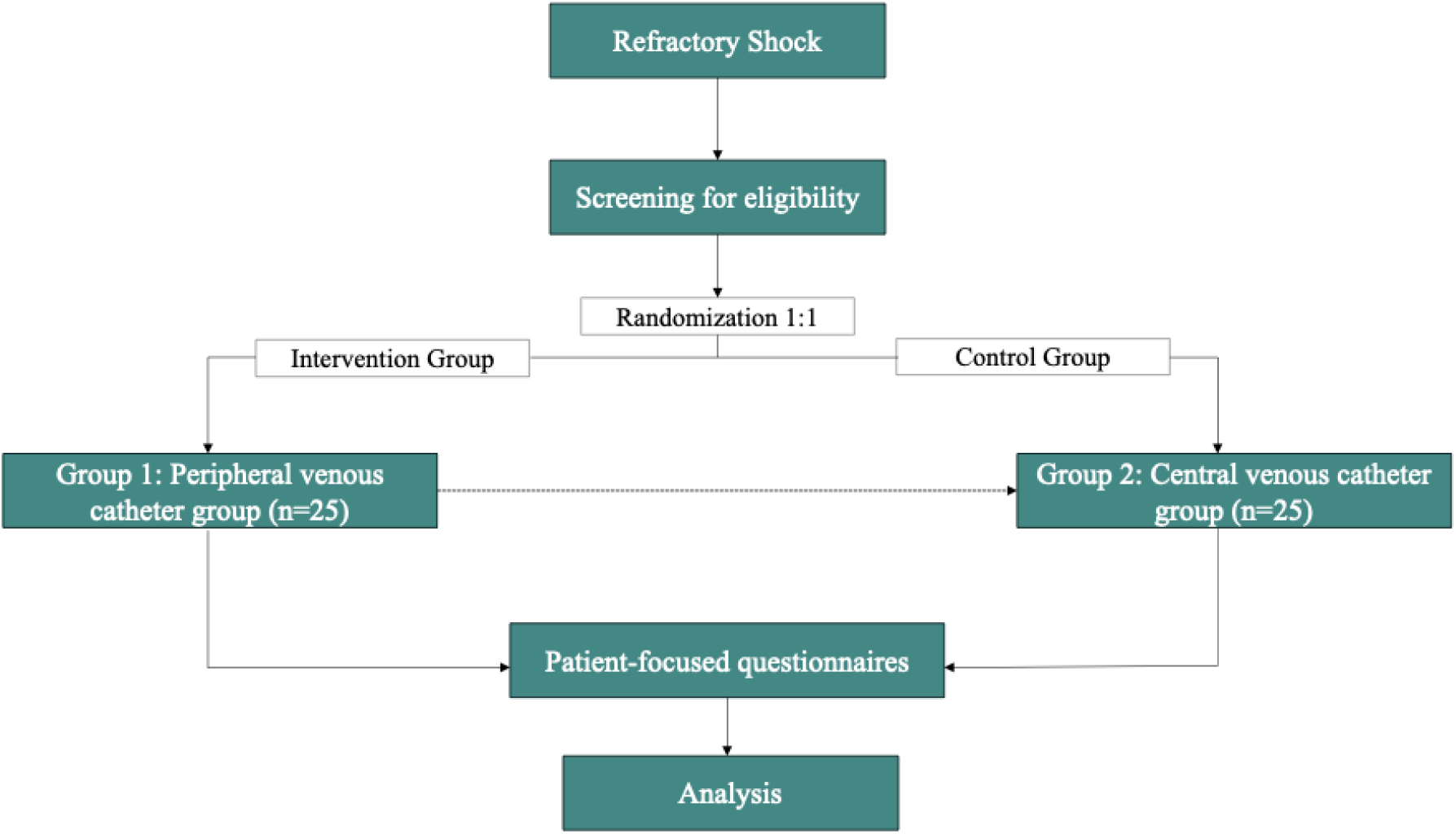
PULSE Study Schema.

### Inclusion criteria

- Adults aged 18 or older.
- No existing central venous catheter or peripherally inserted central catheter.
- The presence of shock requiring vasopressors at the following minimum doses: 0.0625 mcg/kg/min, phenylephrine 0.625 mcg/kg/min, and epinephrine 0.0625 mcg/kg/min. A minimum vasopressor dose requirement will be instituted to justify the central line placement.

### Exclusion criteria

- Urgent need for dialysis requiring placement of a hemodialysis catheter at the time of screening
- More than two vasopressors are necessary to maintain a mean arterial pressure (MAP) of greater than 65 mmHg.
- The placement of a central line is not aligned with the patient’s goals of care.
- Expected death in less than 24 hours.
- Patient has had a vasopressor infusion for more than six hours at the time of screening.
- Require central access for medications other than vasopressors (eg. total parenteral nutrition).

### Crossover criteria

Patients who crossover from the PVC group to the CVC group include the following:

- The maximum vasopressor dose has been reached, as indicated in Table 1 or patient requires more than two vasopressors to maintain a MAP greater than 65.
- The patient has a difficult-to-maintain peripheral venous catheter that requires central access.
- The patient requires central access for medications other than vasopressors
- Clinical team preference for central access.

**Table 1:**
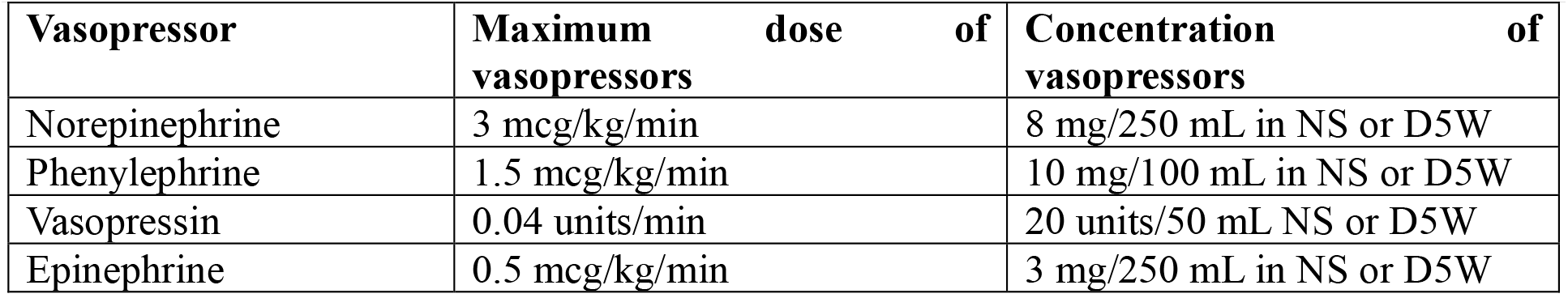
Maximum dose and concentration of peripheral vasopressor agents.

### Waived consent

Our local Health Sciences Research Ethics Board has waived the need for consent, as this study compares the effectiveness of two standard treatments: vasopressor administration via a peripheral venous catheter and a central venous catheter. The peripheral administration of vasopressors is considered safe and low risk, as supported by multiple observational studies. ^5,7,10,11,13,14,17-23^ Patients enrolled will require urgent intervention and will meet all inclusion criteria. In this context, obtaining prior consent would delay time-sensitive treatment and could result in significant harm or death. A debrief will be provided to participants or their surrogate decision-makers (SDMs) if the participant lacks the capacity to consent and is unable to participate in the discussion. During the debrief session, patients and/or SDMs are invited to participate in a brief questionnaire to assess patient-important factors regarding complications that may occur with the placement and/or management of central and peripheral access. The questionnaire guide is found in Supplemental Material 1. Participants and SDMs will be informed of the voluntary nature of the research and their right to withdraw from the study at any time. If withdrawal occurs during the recruitment phase, all associated data will be promptly removed and deleted.

### Screening and Enrollment

Upon initiation of a vasopressor, a member of the clinical care team (including nurses and/or physicians) will inform the research team of any eligible participants. A member of the research team will review the eligibility criteria and the electronic medical record for the following:

- **Renal function:** if renal function is abnormal, the research team will discuss with the clinical care team whether a hemodialysis catheter is needed.
- **Vasopressor dose:** to ensure the minimum dose has been met.
- **Peripheral venous access:** adequate peripheral access will be confirmed with the clinical care team.
- **Research opt-out status:** will be verified via the electronic medical record to ensure the patient has not declined participation in research.

### Randomization

Patients who meet all the inclusion criteria and none of the exclusion criteria will be enrolled in the study. A designated member of the research team will access a password-protected website containing a pre-generated, computerized, randomized list to ensure a concealed 1:1 allocation of study participants to the CVC or PVC group.

### Interventions

**Group 1: Peripheral Venous Catheter Group (intervention group)**

Peripheral vasopressors will be administered exclusively through a PVC inserted in the upper extremities, positioned proximal to the wrist but distal to the antecubital fossa. The following protocol is recommended for safe administration of peripheral vasopressors:

- **The PVC must demonstrate brisk blood return**. If blood return is inadequate, the infusion must be discontinued immediately, and a new catheter site should be established.
- **An 18-gauge or larger PVC is preferred** to ensure adequate flow and reduce the risk of complications.
- **The blood pressure cuff should be placed on the arm opposite** the one used for peripheral vasopressor infusion to avoid issues with catheter patency and monitoring.
- **The catheter site must be inspected five minutes after initiating the infusion and at least hourly thereafter** to monitor for early signs of extravasation or local complications.
- **The maximum duration is 72 consecutive hours** of peripheral vasopressor administration.

The maximum dose and concentration of vasopressor permitted for peripheral administration were defined by prior observational studies and in collaboration with an ICU pharmacist (Table 1).

If extravasation or infiltration occurs, the provider will be immediately notified, and infusion will be switched to an alternate site. The PVC with the infiltrated line will remain in place, the residual medication will be aspirated, and the extravasation site will be outlined. Extravasation kits will be made available to the clinical care team and include subcutaneous needles, instructions for reconstitution and administration of phentolamine.

**Group 2: Central Venous Catheter Group (control group)**

Participants randomized to the control group may have vasopressors administered through a PVC until the clinical care team has placed a central venous catheter. After placement, all vasopressors must be exclusively infused through the central access. The clinical care team has 6 hours to place a CVC once the patient has been recruited into the study.

A standard central-line kit (Pressure injectable Arrow+ard Blue Plus ® Three Lumen CVC) is used. A qualified member of the clinical care team will perform the CVC insertion, ideally under ultrasound guidance to reduce the risk of complications. The clinician performing the procedure will follow a central-line insertion checklist to promote standardization and minimize the risk of infection and other complications. The choice of insertion site will remain at the discretion of the provider. A standardized documentation template for CVC placement, which includes the components of the checklist, is available for use by the clinical care team. A chest x-ray is recommended to ensure proper placement of CVC (i.e. at the junction of the superior vena cava and right atrium) prior to use for CVCs that are placed in the jugular or subclavian veins. Any complications associated with the placement of the CVC will be documented and made available to the research team.

Routine monitoring for complications associated with the maintenance of CVCs will be completed by members of the research team. This will include monitoring for the following:

- Thrombosis
- Infection
- Catheter-migration
- Catheter-occlusion
- Arrythmias

### Patient and public involvement

A significant challenge in designing a clinical trial for peripheral vasopressors is defining severe adverse events, as the complications associated with peripheral venous catheters differ substantially from those of central venous catheters. To address this, questionnaires will be administered to qualitatively explore outcomes important to patients and/or their families, specifically those related to complications arising from the placement and/or maintenance of CVCs and PVCs. The questionnaire was developed in partnership with patient advocates to ensure inclusion of patient-identified priorities and relevant issues. The data obtained from the questionnaires will help to define severe adverse events in a definitive randomized controlled trial.

### Data collection

Data will be de-identified and stored securely in a REDCap® database, which is housed on the Queen’s University Center for Advanced Computing Server. A Master Linking Log will contain only the participant’s record ID and Medical Record Number. The Master Linking Log will be stored separately from the REDCap® database, on a secure university network that is password-protected and restricted to members of the research team. The research team will collect demographics, illness severity, admitting date and diagnosis, access site, number of attempts of access placement, presence of arterial line in PVC group, vasopressor type and maximum vasopressor dose, duration of vasopressor, complications, volume status, ICU and hospital length of stay, discharge and/or transfer date, and in-hospital death if applicable. Information collected in the questionnaire will be transcribed onto the interview guide found in Supplement Material 1, which will be anonymized. This data will be transferred to a REDCap ® database.

Despite these comprehensive safeguards, there is a potential risk of a privacy breach. To prevent a breach, all data will be recorded directly into the REDCap ® database, which can only be accessed by the research team.

### Primary outcomes

The primary outcome will be to assess the feasibility of a peripheral vasopressor protocol, which will include the following:

1. **Recruitment rate** (at least one participant recruited per week or 4 participants per month)
2. **Data capture rate** (≥ 90% of data captured by study participants)
3. **Conversion** rate from peripheral to central access (< 50% of patients initially allocated to the PVC group require CVC placement).

### Secondary outcomes

We will assess the following:

1. **Safety of vasopressors administered via PVC**, namely extravasation rates and phentolamine administration.
2. **Days alive and central-line-free**.
3. **Number of attempts for the placement of either the PVC or CVC prior to starting vasopressors**.
4. **Complication rate** in the central venous catheter group. This will include complications from the insertion of the line (e.g., pneumothorax, erroneous arterial cannulation, and bleeding) and management of the line (e.g., infection, venous thromboembolism).
5. **Volume of vasopressor administration** in both the peripheral venous catheter group and the central venous catheter group. This is because the peripheral administration of vasopressors requires a more dilute concentration and, thus, a higher volume to attain the same dose.
6. **Hospital mortality rate** in both the peripheral venous catheter and central venous catheter groups.
7. **ICU and hospital length of stay** in both the peripheral venous catheter and central venous catheter groups.
8. **Patient-centered important outcomes** regarding complications that can arise in either the CVC or PVC groups.

### Sample size

Given that a feasibility study will be conducted, no formal sample size calculation is needed. Based on monthly admission rates at the intensive care unit, we aim to recruit 50 participants in the trial (25 in the PVC group and 25 in the CVC group). The anticipated timeframe for recruiting participants will be over 12 months.

### Data analysis

The study population will be analyzed using an intention-to-treat analysis. All patients enrolled in the study will be analyzed according to the group to which they were initially randomized, regardless of the intervention they received or adherence to the protocol.

For the pilot study, the primary outcomes were calculated with 95% confidence intervals. For demographics, we used counts (percentages) for categorical variables, and for secondary outcomes, we used means (standard deviations [SD]) for normally distributed variables or medians (interquartile ranges [IQR]) for skewed continuous variables. As this was a feasibility study, missing data were not imputed, and analyses were based solely on observed data.

For patient-centred outcomes, the questionnaire will be analyzed for common themes, and the ranking of complications will be described using descriptive statistics with 95% confidence intervals.

## Supporting information

Patient Questionnaire

The PULSE Protocol

## Data Availability

All data produced in the present study are available upon reasonable request to the authors

## ETHICS AND DISSEMINATION

Approval for the study has been obtained from the Queen’s University Ethics Board, REB approval number: 6042888. This feasibility study aims to generate data to inform the development of a protocol for the peripheral administration of vasopressors, thereby informing the design and implementation of a definitive trial. We plan to present the study’s findings at local, national, and international scientific conferences and meetings. We will also submit the findings to a peer-reviewed journal to disseminate the results to the broader critical care community.

## IMPLICATIONS

The data collected from this study will provide valuable information to conduct a larger randomized controlled trial, which will help fill the knowledge gap necessary to ensure that peripheral administration of vasopressors is safe. Future research should focus on patient-centred outcomes and on deriving a protocol that is generally applicable across many institutions. This patient input will help to define severe adverse events, as the complications associated with peripheral venous catheters differ substantially from those of central venous catheters. Most importantly, we must ensure that healthcare providers caring for critically ill patients prioritize obtaining appropriate evidence-based access rather than delaying treatment for unnecessary procedures. ^24^

