## Supplementary material for "The Peripheral Use of Low-dose Vasopressors for Safety and Efficacy (PULSE) in the intensive care unit: a prospective, unblinded feasibility study protocol": Patient Questionnaire

**Supplementary Material 1**

### QUESTIONNAIRE GUIDE

**DEFINITIONS/TERMS:**

| | |
|---|---|
| **Central venous catheter or central line** | A thin, flexible tube that doctors place into a large vein, usually in the neck, chest, or groin. |
| **Peripheral venous catheter or Peripheral IV** | A small, short plastic tube that's placed into a vein, usually in your hand or arm. It's referred to as peripheral because it goes into a smaller vein near the surface of your body |
| **Collapsed lung** | When air leaks into the space between the lung and the chest wall. This air pushes on the lung and makes it collapse. It can cause sudden chest pain and trouble breathing. It can happen after an injury or medical procedure. Oftentimes, a chest tube, which is a flexible plastic tube that doctors insert through the chest wall is needed to help remove the air. |
| **Vasopressors** | A drug that helps raise your blood pressure. It's often used when someone's blood pressure is dangerously low, like during a severe infection or shock, to help keep blood flowing to important organs. |
| **Extravasation** | When a drug or fluid that's supposed to go into your vein accidentally leaks into the surrounding tissue. This can cause pain, swelling, or even tissue damage, especially if the drug is strong or irritating. |

#### QUESTIONS

**1. To give some medicines, a small tube is placed into a vein. To do this, a needle is inserted through the skin into the vein, allowing a small tube to be placed. Once the tube is in the right spot, the needle is removed, and the tube remains in place.**

a. A central line is a type of tube that is inserted into a deeper vein, requiring a larger needle. To help alleviate the pain associated with a central line, a numbing medicine is injected under the skin using a small needle. This procedure requires a skilled doctor and often an ultrasound to prevent complications. There is a risk of infection, so it must be done under sterile conditions. A central line can stay in for a long time, which means fewer needle pokes later.

b. For a peripheral IV, the vein is closer to the skin, and a smaller needle is used to insert the tube. The peripheral IV may not last as long as a central line; therefore, some patients may require replacement of the peripheral IV, which would result in more needle pokes.

Of the following, rank the following from most important to least important (1=most important, 4=least important):

- ☐ The number of needle sticks needed
- ☐ How long can the tube stay in place
- ☐ How complicated the procedure is
- ☐ How much it hurts

**2. Some complications can occur with both central lines and peripheral IVs; some are more serious than others.**

a. Central line complications: collapsed lung, the tube being placed in an artery rather than a vein and infection.
   i. A collapsed lung requires placement of a large tube in the chest to remove the air and allow the lung to expand. A collapsed lung can lead to death and requires urgent intervention.
   ii. If the tube is placed in the artery, it requires a surgeon or specialist to remove the line.
   iii. If you become infected secondary to the central line, a course of antibiotics may be required.

b. Peripheral IV complications include extravasation (the leakage of medicine into the surrounding tissue, as the tube is no longer in the vein), difficulty maintaining peripheral IVs (the IV is no longer functioning or falls out), and skin infection around the peripheral IV site.
   i. In severe cases, extravasation can cause tissue damage and may require surgical removal of affected tissue.
   ii. Difficult-to-maintain peripheral IVs need to be removed, and a new peripheral IV is placed.
   iii. Skin infection: Most cases resolve with the removal of the peripheral IV, and in some cases, a course of antibiotics may be required.

Do you think a difficult-to-maintain peripheral IV is as severe as a collapsed lung?

- ☐ Yes
- ☐ No
- ☐ Not sure

If both a peripheral IV and a central line were appropriate in the management of your care, after hearing the complications that can occur with both, which would you prefer:

- ☐ Peripheral IV
- ☐ Central line
- ☐ Not sure

Of the following, rank the following from most important to least important (1=most important, 3=least important):

- ☐ Severity of complication
- ☐ A complication that can lead to disfigurement
- ☐ Whether the complication increases the number of days you need to stay in the hospital

**3. When in the hospital and particularly in the intensive care unit, blood work is ordered at least every day and sometimes multiple times in a day. After a central line has been placed, blood can be drawn from this line. This means that the nurses do not need to poke your arm for blood work.**

**Sometimes, peripheral IVs can be used for blood work, but this increases the risk that the peripheral IV may no longer work; therefore, most patients need to undergo a needle stick for blood work.**

Does the need for frequent blood work impact whether or not you would prefer a central line or a peripheral IV?

- ☐ Yes
- ☐ No
- ☐ Not sure
