## Supplementary material for "The Peripheral Use of Low-dose Vasopressors for Safety and Efficacy (PULSE) in the intensive care unit: a prospective, unblinded feasibility study protocol": The PULSE Protocol

### The PULSE Study

---

**Principle Investigator**: Dr. J. Gordon Boyd, MD PhD FRCPC

**MSc Candidate:** Jessica Wiseman

**Research Team:** Dr. Stephanie Sibley, Dr. Santiago Perez, Maikel Mekhaeil, Tracy Boyd, Miranda Hunt, Michaela Hanley, Brianna Grant

### TABLE OF CONTENTS

**1. Introduction**
1.1 Background
1.2 Rationale and Clinical Significance

**2. Study Objectives**
2.1 Primary Objective
2.2 Secondary Objectives

**3. Study Details**
3.1 Study Design
3.2 Study Schema
3.3 Intervention
3.4 Primary Outcomes
3.5 Secondary Outcomes
3.6 Peripheral Vasopressor Protocol
3.7 Extravasation Protocol
3.8 Central Line Placement Protocol
3.9 Patient-centered Outcomes

**4. Study Population**
4.1 Eligibility Criteria
4.2 Exclusion Criteria
4.3 Crossover Criteria
4.4 Recruitment

**5. Data Collection, Management and Statistical Considerations**
5.1 Data Collection
5.2 Data Analysis
5.3 Sample Size Estimate

**6. Ethical Considerations**
6.1 Research Ethics and Dissemination
6.2 Consent

**7. Knowledge Translations**

**8. Appendix**
8.1 Extravasation Protocol
8.2 Template for Central-line Placement
8.3 Interview Guide for Patient-Centred Important Outcomes

**9. References**

### 1. INTRODUCTION

#### 1.1 Background

In the intensive care unit (ICU), vasopressors are administered when fluid resuscitation alone cannot maintain adequate blood pressure in patients who present with shock[1], which is a life-threatening condition characterized by inadequate tissue perfusion. The central venous catheter (CVC) or central line has been the preferred route of administration for vasopressors, based on historical data, due to the risk of vasopressor extravasation through peripheral venous catheters (PVCs). [2] With the introduction of norepinephrine to the market in the 1950s, the incidence of extravasation associated with its peripheral administration ranged from 46% to 60%. [3] However, advances in venous catheter access have significantly reduced this risk, with current extravasation rates now ranging from 1.8% to 3.4%, as reported in two recent systematic reviews. [2,4]

The placement of a CVC requires a skilled clinician and, oftentimes, ultrasound guidance to prevent complications such as pneumothorax, bleeding, or inadvertent arterial cannulation. Furthermore, complications related to the maintenance of CVCs include deep vein thrombosis (DVT) and central line-associated bloodstream infections (CLABSIs). These recognized complications may occur in more than 15% of patients. [5,6] Therefore, there is a growing interest in peripheral vasopressor use for two reasons: 1) to expedite vasopressor initiation in patients with shock and 2) to avoid CVC placement and its potential complications.[7] Many clinicians initiate peripheral vasopressors in patients presenting with refractory shock, leading to a shorter vasopressor administration time than central venous access [8-10]. However, the continued use of peripheral vasopressors remains rare, as most patients eventually require central access. [8]

#### 1.2 Rationale & Clinical Significance

The current evidence on the use of peripheral vasopressors is primarily based on single-centre observational studies. The inconsistency in peripheral vasopressor administration, monitoring of peripheral lines, and variations in vasopressor concentrations, doses, and duration of use contribute to hospitals' reluctance to adopt peripheral vasopressors and to develop best practices.[8,18] Additionally, there are no universal guidelines for the transition threshold for central-line placement, with only weak recommendations from the Surviving Sepsis Guidelines suggesting that vasopressors should be infused through a central line as soon as possible.[19] Therefore, many suggest that PVCs should be viewed as a bridge to central access until more reliable evidence becomes available. [20]

A randomized controlled trial is needed to provide high-quality evidence to improve the future use of peripheral vasopressors, which could positively impact patient care. Future research should focus on patient-centred outcomes and on deriving a protocol that is generally applicable across many institutions. The patients who would benefit the most are those who present with mild shock and are expected to be on low-dose vasopressors for short durations.

### 2. STUDY OBJECTIVES

#### 2.1 Primary Objectives

To determine the feasibility of a randomized controlled trial for the implementation of a peripheral vasopressor protocol. The following factors will be assessed:

1. Recruitment rate (measured as the proportion of eligible patients randomized into the study).
2. Data capture rate (measured as the percentage of data elements obtained for each participant).
3. Conversion rate to central access (measured as the number of participants recruited into the PVC arm who require CVC placement).

##### 2.2 Secondary Objectives

1. Assess the safety of peripheral vasopressors.
2. Assess alive and central-line-free days.
3. Assess the number of attempts in the placement of the peripheral venous catheter and the central venous catheter attempts.
4. Assess the rate of complications.
5. Determine the amount of vasopressor volume.
6. Assess the 30-day mortality rate.
7. Compare ICU and hospital length of stay.
8. Assess patient-centred important outcomes.

#### 3. STUDY DETAILS

##### 3.1 Study design

The proposed study design is a prospective, unblinded feasibility study. It will be unblinded because a central line will be easily noticeable. The trial is registered with clinicaltrials.gov (trial registration number: NCT06920173).

##### 3.2 Study schema

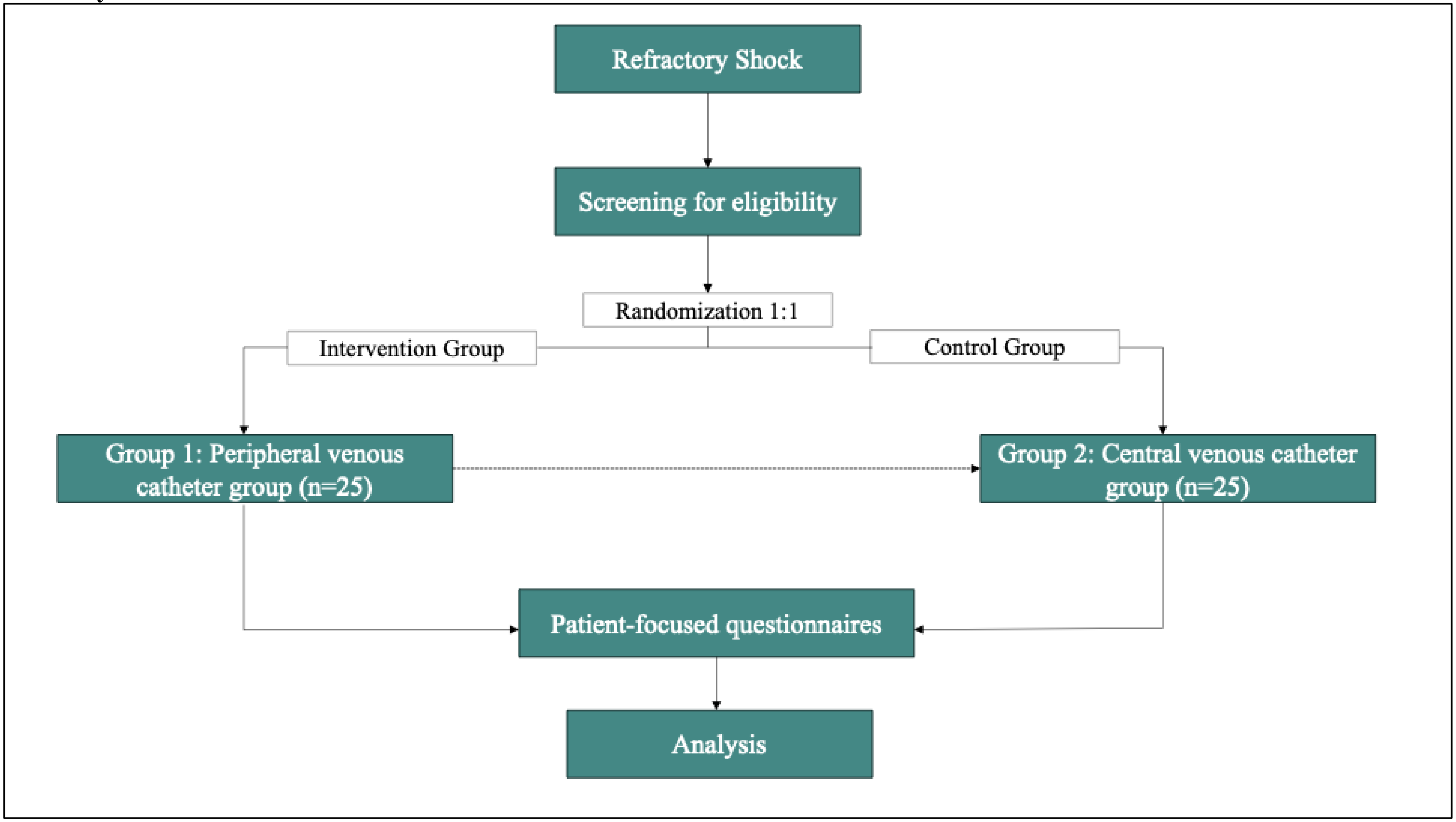


**Figure 1. Study schema**

##### 3.3 Intervention

Upon initiation of a vasopressor, a member of the care team will notify the research team to assess study eligibility. If eligible, a designated member of the research team will access a password-protected website containing a pre-generated, computerized, randomized list to ensure a concealed 1:1 allocation of study participants to the CVC or PVC group.

**Group 1: Peripheral Venous Catheter Group (intervention group)**

Peripheral vasopressors will be administered exclusively through a PVC inserted in the upper extremities, positioned proximal to the wrist but distal to the antecubital fossa. The following protocol is recommended for safe administration of peripheral vasopressors:

- **The PVC must demonstrate brisk blood return.** If blood return is inadequate, the infusion must be discontinued immediately, and a new catheter site should be established.
- **An 18-gauge or larger PVC is preferred** to ensure adequate flow and reduce the risk of complications.
- **The blood pressure cuff should be placed on the arm opposite** the one used for peripheral vasopressor infusion to avoid issues with catheter patency and monitoring.
- **The catheter site must be inspected five minutes after initiating the infusion and at least hourly thereafter** to monitor for early signs of extravasation or local complications.
- **The maximum duration is 72 consecutive hours** of peripheral vasopressor administration.

The maximum dose and concentration of vasopressor permitted for peripheral administration were defined by prior observational studies and in collaboration with lead ICU pharmacist Brianna Grant (PharmD, RPh, ACPR) at KHSC. Table 1 provides the maximum doses and concentrations of each vasopressor used in the PVC group.

| Vasopressor | Maximum dose of vasopressors | Concentration of vasopressors |
|---|---|---|
| Norepinephrine | 3 mcg/kg/min | 8 mg/250 mL in NS or D5W |
| Phenylephrine | 1.5 mcg/kg/min | 10 mg/100 mL in NS or D5W |
| Vasopressin | 0.04 units/min | 20 units/50 mL NS or D5W |
| Epinephrine | 0.5 mcg/kg/min | 3 mg/250 mL in NS or D5W |

**Table 1:** Maximum dose and concentration of peripheral vasopressor agents.

If extravasation or infiltration occurs, the provider will be immediately notified, and infusion will be switched to an alternate site. The PVC with the infiltrated line will remain in place, the residual medication will be aspirated, and the extravasation site will be outlined. Extravasation kits including subcutaneous needles, instructions for reconstitution and administration of phentolamine will be made available to the clinical care team. Appendix 8.1 outlines the full protocol for extravasation.

| **Group 2: Central Venous Catheter Group (control group)** |
|---|
| Participants randomized to the control group may have vasopressors administered through a PVC until the clinical care team has placed a central venous catheter. After placement, all vasopressors must be exclusively infused through the central access. The clinical care team has 6 hours to place a CVC once the patient has been recruited into the study.<br><br>The standard central-line kit used at KGH is the Pressure injectable Arrow+ard Blue Plus ® Three Lumen CVC. A qualified member of the clinical care team (including junior residents under the supervision of an attending physician or critical care fellow) will perform the CVC insertion, ideally under ultrasound guidance to reduce the risk of complications. The clinician performing the procedure will follow a central-line insertion checklist to promote standardization and minimize the risk of infection and other complications. The choice of insertion site will remain at the discretion of the provider. A standardized documentation template for CVC placement (Appendix 8.2), which includes the components of the checklist, is available for use by the clinical care team. A chest x-ray is recommended to ensure proper placement of CVC (i.e. at the junction of the superior vena cava and right atrium) prior to use. Any complications associated with the placement of the CVC will be documented and made available to the research team.<br><br>Routine monitoring for complications associated with the maintenance of CVCs will be completed by members of the research team. This will include monitoring for the following:<br>• Thrombosis<br>• Infection<br>• Catheter-migration<br>• Catheter-occlusion<br>• Arrythmias<br>• Bleeding |

##### 3.4 Primary Outcomes

The primary outcome will be to assess the feasibility of a peripheral vasopressor protocol, which will include the following:

1. **Recruitment rate** (at least one participant recruited per week or 4 participants per month)
2. **Data capture rate** ($\geq$ 90% of data captured by study participants)
3. **Evaluate the rate of conversion** from peripheral to central access, aiming for no more than 50% of patients initially allocated to the PVC group to require CVC placement.

##### 3.5 Secondary Outcomes:

1. **Assess the safety of peripheral vasopressors,** including capturing extravasation rates and determining the need for phentolamine administration.
2. **Assess alive and central-line-free days.**
3. **Assess the number of attempts in the placement of the peripheral venous catheter and the central venous catheter attempts** required to start vasopressors.
4. **Assess the rate of complications** in the central venous catheter group. This will include complications from line insertion (e.g., pneumothorax, erroneous arterial cannulation, and bleeding) and from line management (e.g., infection, venous thromboembolism).

5. **Determine the amount of vasopressor volume** in both the peripheral venous catheter group and the central venous catheter group.
6. **Assess the mortality rate** in both the peripheral venous catheter and central venous catheter groups.
7. **Compare ICU and hospital length of stay** in both the peripheral venous catheter and central venous catheter groups.
8. **Assess patient-centred important outcomes** related to complications that can arise in either the CVC or the PVC groups.

##### 3.8 Patient-centred outcomes

A significant challenge in designing a clinical trial for peripheral vasopressors is defining severe adverse events, as the complications associated with peripheral venous catheters differ substantially from those of central venous catheters. To address this, focused questionnaires will be conducted to qualitatively explore outcomes that are important to patients and/or their families, specifically those related to complications arising from the placement and/or maintenance of CVCs and PVCs. Participants and/or family members will be recruited during the debriefing process, as consent for this study has been waived. The focused questionnaire will be transcribed using the interview guide found in Appendix 8.3. The questionnaire was adapted with the input of patient advisers at Kingston Health Sciences Center.

#### 4. STUDY POPULATION

##### 4.1 Eligibility Criteria

- Adults aged 18 or older.
- No existing central venous catheter or peripherally inserted central catheter patients who are receiving vasopressors through a peripheral venous catheter.
- The presence of shock requiring vasopressors at the following minimum doses: norepinephrine five mcg/min, phenylephrine 50 mcg/min, epinephrine five mcg/min, dobutamine five mcg/kg/min. A minimum vasopressor dose requirement will be instituted to justify the placement of a central line.

##### 4.2 Exclusion Criteria

- Urgent need for dialysis requiring placement of a hemodialysis catheter at the time of screening
- The patient already has central access in place (i.e., central venous catheter or peripherally inserted central catheter).
- More than 2 vasopressors are required to maintain a mean arterial pressure (MAP) greater than 65 mmHg.
- The placement of a central line is not aligned with the patient's goals of care.
- Suspected death in less than 24 hours.
- The patient has had a vasopressor infusion for more than six hours at the time of screening.
- Require central access for medications other than vasopressors (e.g., total parenteral nutrition).

##### 4.3 Crossover Criteria

Patients who crossover from the PVC group to the CVC group include the following:

- The maximum vasopressor dose has been reached, as indicated in Table 1

- The patient has a difficult-to-maintain peripheral venous catheter, necessitating central access.
- The patient requires central access for medications other than vasopressors
- Clinical team preference for central access.

##### 4.4 Recruitment

Upon initiation of a vasopressor, a member of the clinical care team (including nurses and/or physicians) will inform the research team of any eligible participants. A member of the research team will review the eligibility criteria and the electronic medical record for the following:

- **Renal function:** if renal function is abnormal, the research team will discuss with the clinical care team whether a hemodialysis catheter is needed.
- **Vasopressor dose:** ensure the minimum dose is met.
- **Peripheral venous access:** Adequate peripheral access will be confirmed with the clinical care team.
- **Research opt-out status:** will be verified via the electronic medical record to ensure the patient has not declined participation in research.

This study has received a waiver of informed consent; however, a debrief will be provided to participants or their surrogate decision-makers if the participant is unable to engage in the discussion. Additional details regarding the waived consent and debriefing process are outlined in Section 6, "Ethical Considerations."

#### 5. DATA COLLECTION, MANAGEMENT AND STATISTICAL CONSIDERATIONS

##### 5.1 Data Collection and Management

Data will be de-identified and stored securely in a REDCap® database, which is housed on the Queen's University Center for Advanced Computing Server. A Master Linking Log will contain only the participant's record ID and Medical Record Number. The Master Linking Log will be stored separately from the REDCap® database on the Kingston Health Sciences Center network, which is password-protected and restricted to members of the research team. Data collected will include demographics, completion of debrief, illness severity, admitting date and diagnosis, access site, number of attempts of access placement, presence of arterial line in PVC group, vasopressor type and maximum vasopressor dose, duration of vasopressor, complications, volume status, ICU and hospital length of stay, discharge and/or transfer date, in-hospital death if applicable.

All transcribed data from the interview guide for patient-centred outcomes will be stored in a REDCap® database accessible only to members of the research team. There will be no identifiable information on the paper copy of the interview guide. Data will be retained for five years following the completion of the study, in accordance with Queen's University policy, and then securely destroyed.

Despite these comprehensive safeguards, there is a potential risk of a privacy breach. In the event of an unauthorized disclosure of personal information, immediate steps will be taken to contain the breach and recover the data. The KHSC Privacy Office, Queen's University Privacy Office, and the Research Ethics Board (REB) will be promptly notified. Any additional actions recommended by these bodies will be followed in full.

##### 5.2 Data Analysis

The study population will be analyzed using an intention-to-treat analysis. All patients enrolled in the study will be analyzed according to the group to which they were initially randomized, regardless of the intervention they received or adherence to the protocol.

For the pilot study, the primary outcomes were calculated with 95% confidence intervals. For demographics, we used counts (percentages) for categorical variables, and for secondary outcomes, we used means (standard deviations [SD]) for normally distributed variables or medians (interquartile ranges [IQR]) for skewed continuous variables. As this was a feasibility study, missing data were not imputed, and analyses were based solely on observed data.

Alive and central line–free days were defined as the number of days within the 28-day observation period during which a patient was alive and without a central venous catheter. Patients who died before day 28 were assigned zero alive and central line–free days. Any calendar day with a CVC present for any duration was counted as a CVC day.

For patient-centred outcomes, the questionnaire will be analyzed for common themes, and the ranking of complications will be described using descriptive statistics with 95% confidence intervals.

##### 5.3 Sample Size Estimate

Given that a feasibility study will be conducted, there is no formal calculation for sample size. Based on monthly intensive care unit admission rates at Kingston General Hospital, we aim to recruit 50 participants to the study (25 in the PVC group and 25 in the CVC group).

#### 6. ETHICAL CONSIDERATIONS

##### 6.1 Research Ethics

Approval for the study has been obtained from the Queen's University Ethics Board. In compliance with the Queen's University Tri-Council Policy Statement, the research team will have completed the ethics training (TCPS2 CORE).

##### 6.2 Waived Consent

Consent will be waived for this study, which compares the effectiveness of two accepted treatments: vasopressor administration via a peripheral venous catheter and a central venous catheter. The use of peripheral vasopressors is considered safe and low risk, as supported by multiple observational studies. [1,6,8,12,14,16,17,23-28] Patients enrolled will require urgent intervention and will meet all inclusion criteria. In this context, obtaining prior consent would delay time-sensitive treatment and could result in significant harm or death. Any modification to the consent process will be documented.

Following enrollment, participants or their substitute decision-makers (SDMs), if the participant is unable to communicate (e.g., due to intubation, sedation, or delirium), will undergo a debriefing session as soon as feasible. During this session, they will be invited to participate in a focused interview to identify patient-centred outcomes relevant to the choice of vascular access (CVC vs PVC). If they agree, they will be asked a series of pre-determined questions outlined in Appendix 8.3.

Participants and SDMs will be informed of their right to withdraw from the study at any time. If withdrawal occurs during the recruitment phase, all associated data will be promptly removed and deleted.

#### 7. KNOWLEDGE TRANSLATION

This feasibility study aims to generate data to inform the development of a peripheral vasopressor protocol for conducting larger clinical trials. We plan to present the study's findings at local, national, and international scientific conferences and meetings. We will also consider submitting the findings of a peer-reviewed manuscript to a relevant journal to disseminate the results to the broader critical care community.

#### 8. APPENDIX

##### 8.1 Extravasation Protocol

1. **Prevention of extravasation**
    - The use of superficial veins in the forearm is preferred, and flexion points should be avoided.
    - Avoid multiple attempts while attempting venous access.
    - Assess vein integrity before initiating a peripheral vasopressor and ensure the peripheral venous catheter (PVC) has a good blood return.
    - Monitor PVC for early signs and symptoms of infiltration/extravasation (Table 1: Signs and symptoms of extravasation)
    - Advise patient to notify if the following symptoms occur such as stinging, burning or patient at the PVC site.

2. **General treatment of extravasation or infiltration**
    - Stop and disconnect the infusion from the PVC.
    - Aspirate as much of the medication and then remove the PVC.
    - Mark the extravasation area, if possible, to allow for monitoring of injury
    - Grade the infiltration using Table 1: Signs and Symptoms of Extravasation
    - Elevate the affected limb if able to reduce swelling
    - Notify the provider and obtain order for further treatment.
    - See Table 2: Pharmacologic Treatment for Management of Extravasation
    - Prior to any subcutaneous or intradermal treatment, cleanse the skin with an alcohol wipe
    - Observe and document patient response to treatment in patient's medical record

3. **If ulceration occurs following extravasation**
    - Notify provider and seek advice from a surgeon

**Table 1: Signs and Symptoms of Extravasation**

| **Signs** | **Symptoms** |
|---|---|
| Swelling | Tightness |
| Redness | Burning |
| Blanching | Aching |
| Unexplained reduced IV flow rate | Tingling sensation |
| Lack of blood return | Itchiness |

**Directions for reconstitution of phentolamine:**
5 mg vial diluted with 10 mL of NS to achieve a concentration of 0.5 mg/mL. Then take small aliquots (0.2 mL) and administer SC. The total volume to administer depends on the size of the extravasation. (from Up to Date and our parenteral manual). Administer as soon as possible and within 12 hours of extravasation. May readminister in 60 minutes if patient remains symptomatic. If an IV catheter remains in place, recommend administering the initial dose intravenously through the catheter.

**Table 2: Pharmacological Treatment for Management of Extravasation**

| Drug | Pharmacologic Management | Compress Temperature | Comments |
|---|---|---|---|
| Dobutamine<br><br>OR<br><br>Dopamine<br><br>OR<br><br>Epinephrine<br><br>Or<br><br>Norepinephrine | **Preferred:**<br>Phentolamine<br><br>**Alternate:** Topical Nitroglycerin 2% | Warm | May repeat if no improvement in 1 hour.<br><br>Monitor BP and HR<br>Monitor BP, HR, and respiratory rate<br><br>Phentolamine- best within<br>12 hours.<br><br>Terbutaline- best within 1-2 hours. |
| Phenylephrine | **Preferred:**<br>Phentolamine<br><br>**Alternate:**<br>Topical Nitroglycerin 2% | Warm | May repeat if no improvement in 1 hour.<br><br>Monitor BP and HR<br><br>Phentolamine- best within<br>12 hours. |
| Vasopressin | **Preferred:**<br>Topical Nitroglycerin 2%<br><br>**Alternate:**<br>Phentolamine | Warm | May repeat if no improvement in 1 hour.<br><br>Monitor BP and HR<br><br>Phentolamine- best within<br>12 hours |

##### 8.2 Central Line Procedure Note/Checklist

**Procedure Note: Central Line Placement**
**Kingston Health Sciences Center**

Indications: _▼

The procedure is: _▼

Insertion Site: _▼

Number of attempts: _▼

The coagulation profile and platelet count were reviewed prior to placement of the central line. The patient's ID (full name, date of birth and/or MRN) were verified. The insertion site was assessed/marked, and the patient was correctly positioned for the procedure. The equipment was assembled, and supplies verified. Prior to placement, it was confirmed that all persons in the room had cleansed their hands.

Provider and assistant (if present), wore gloves, hat, mask and sterile gown. All persons in the room wore masks. The patient's skin was prepped with chlorhexidine 2%/70% alcohol, allowing sufficient time to dry (~2 minutes). A large sterile drape was used to cover the patient. Local anesthesia was provided with _▼. Sedation provided with _▼. Ultrasound guidance is used to cannulate the vein and confirm venous placement. Each lumen has good blood return and is flushed with sterile water. All lumens are capped. The central line was sutured in place, and a sterile dressing was placed. A chest x-ray was ordered to confirm placement.

**Comments and/or complications during insertion:**

##### 8.3 Questionnaire for Patient-Centred Important Outcomes

**SETTING**

This questionnaire/interview guide will be administered during the debriefing session, after the patient has been randomized into the study, into either the central venous catheter arm or the peripheral venous catheter arm. A member of the research team will conduct the debriefing session/interview.

- ☐ Yes
- ☐ No
- ☐ Not sure
